# Cohort Profile: a Prospective Household cohort study of Influenza, Respiratory Syncytial virus, and other respiratory pathogens community burden and Transmission dynamics in South Africa (PHIRST), 2016-2018

**DOI:** 10.1101/2021.01.06.21249313

**Authors:** Cheryl Cohen, Meredith L. McMorrow, Neil A. Martinson, Kathleen Kahn, Florette K. Treurnicht, Jocelyn Moyes, Thulisa Mkhencele, Orienka Hellferscee, Limakatso Lebina, Matebejane Moroe, Katlego Motlhaoleng, F. Xavier Gómez-Olivé, Ryan Wagner, Stephen Tollman, Floidy Wafawanaka, Sizzy Ngobeni, Jackie Kleynhans, Azwifari Mathunjwa, Amelia Buys, Lorens Maake, Nicole Wolter, Maimuna Carrim, Stuart Piketh, Brigitte Language, Angela Mathee, Anne von Gottberg, Stefano Tempia, for the PHIRST group

**Affiliations:** Centre for Respiratory Diseases and Meningitis, National Institute for Communicable Diseases of the National Health Laboratory Service, Johannesburg, South Africa; School of Public Health, Faculty of Health Sciences, University of the Witwatersrand, Johannesburg, South Africa; Influenza Division, Centers for Disease Control and Prevention, Atlanta, Georgia, United States of America; Influenza Program, Centers for Disease Control and Prevention, Pretoria, South Africa; United States Public Health Service, Rockville, Maryland, United States of America; Perinatal HIV Research Unit, Medical Research Council (MRC) Soweto Matlosana Collaborating Centre for HIV/AIDS and Tuberculosis, South Africa; Johns Hopkins University Center for Tuberculosis Research, Baltimore, Maryland, United States of America; Department of Science and Technology/National Research Foundation Centre of Excellence for Biomedical Tuberculosis Research, University of the Witwatersrand, Johannesburg, South Africa; MRC/Wits Rural Public Health and Health Transitions Research Unit (Agincourt), School of Public Health, Faculty of Health Sciences, University of the Witwatersrand, Johannesburg, South Africa; Division of Medical Virology, National Health Laboratory Service, Charlotte Maxeke Johannesburg Academic Hospital, Johannesburg, South Africa; School of Pathology, Faculty of Health Sciences, University of the Witwatersrand, Johannesburg, South Africa; Climatology Research Group, Unit for Environmental Science and Management, School of Geo and Spatial Science, North-West University, Potchefstroom, South Africa; Environment and Health Research Unit, South African Medical Research Council, Johannesburg, South Africa; Environmental Health Department, Faculty of Health Sciences, University of Johannesburg, Johannesburg, South Africa; MassGenics, Duluth, GA, United States of America

**Keywords:** cohort profile, influenza, respiratory syncytial virus, transmission, burden, South Africa

## Abstract

**Purpose:** The PHIRST study (Prospective Household cohort study of Influenza, Respiratory Syncytial virus, and other respiratory pathogens community burden and Transmission dynamics in South Africa) aimed to estimate the community burden of influenza and respiratory syncytial virus (RSV) including the incidence of infection, symptomatic fraction, and disease severity, and to assess household transmission. We further aimed to estimate the impact of HIV infection and age on disease burden and transmission, and to assess the burden of *Bordetella pertussis* and *Streptococcus pneumoniae*.

**Participants:** We enrolled 1684 individuals in 327 randomly selected households in two sites (rural Agincourt subdistrict, Mpumalanga Province and urban Jouberton Township, North West Province) over 3 consecutive influenza and RSV seasons. A new cohort of households was enrolled each year. Eligible households included those with >2 household members where ≥80% of household members provided consent (and assent for children aged 7-17 years). Enrolled household members were sampled with nasopharyngeal swabs twice weekly during the RSV and influenza seasons of the year of enrolment. Serology samples were collected at enrolment and before and after the influenza season annually.

**Findings to date:** There were 122,113 potential individual follow-up visits over the 3 years, and participants were interviewed for 105,783 (87%) of these. Out of 105,683 nasopharyngeal swabs from follow-up visits, 1,258 (1%), 1,026 (1%), 273 (<1%), 38,829 (37%) tested positive on PCR for influenza viruses, respiratory syncytial virus, pertussis and pneumococcus respectively.

**Future plans:** Future planned analyses include analysis of influenza serology results and RSV burden and transmission. Households enrolled in the PHIRST study during 2016-2018 were eligible for inclusion in a study of SARS-CoV-2 transmission initiated in July 2020. This study uses similar testing frequency and household selection methods to assess the community burden of SARS-CoV-2 infection and the role of asymptomatic infection in virus transmission.

**Registration:** Clinical trials.gov NCT02519803

**Article summary:** *Strengths and limitations of this study:*

- PHIRST was conducted in urban and rural African settings with high HIV prevalence, allowing assessment of the effect of HIV on community burden and transmission dynamics of respiratory pathogens.
- Households were selected randomly to provide a representative sample of the community. Twice weekly sampling from each cohort of individuals for 6-10 months irrespective of symptoms allows estimation of community burden, household secondary infection risk, and serial interval including asymptomatic or paucisymptomatic episodes.
- Polymerase chain reaction testing of >100,000 nasopharyngeal swab samples for multiple pathogens (influenza, respiratory syncytial virus, pertussis and *Streptoccocus pneumonia*) allows detailed examination of disease burden and transmission and pathogen interactions
- PHIRST was not powered to assess severe outcomes (i.e. hospitalisation and death).
- We only examined four pathogens, but other micro-organisms may be important. Samples have been stored which could allow us to implement broader multi-pathogen testing in the future.

## Introduction

In 2015, lower respiratory tract infections caused an estimated 2.7 million deaths globally.^1^ Among children aged <5 years, the highest mortality rates are in sub-Saharan Africa where the HIV-epidemic has increased morbidity of severe pneumonia. Influenza, respiratory syncytial virus (RSV), pertussis, and pneumococcus are among the leading causes of pneumonia globally.^2–5^

Approximately 30% of influenza and RSV transmission is estimated to occur within households.^6,7^ Data on community burden and transmission of respiratory pathogens are important to guide vaccination strategies such as reduced pneumococcal conjugate vaccine dose schedules,^8^ optimal timing of booster doses^9^ and vaccinating community transmitters.^10,11^ Illness episodes in the community may be associated with substantial community impact including absenteeism from school or work and loss of income.^12^

The PHIRST study aimed to estimate the community burden of influenza and RSV (including the incidence of infection and symptomatic fraction) and to assess household transmission of influenza and RSV (Supplementary table 1). Secondary objectives included describing the community burden and transmission of *Streptococcus pneumoniae* and *Bordetella pertussis*, estimating the impact of HIV infection and age on disease burden, estimating rates of tuberculosis infection and transmission, and investigating the interaction between respiratory viruses and bacteria. We also aimed to evaluate the role of asymptomatic influenza and RSV infection in household transmission.

## Cohort description

### Study population and household eligibility criteria

A prospective cohort study of randomly selected households in South Africa, was conducted in a rural and an urban site, each with established surveillance for pneumonia and influenza-like illness (Figure 1).^13,14^ The rural site in Mpumalanga Province (Agincourt subdistrict) is part of a health and socio-demographic surveillance system (HDSS), including approximately 116,000 people in 31 contiguous villages.^15,16^ Approximately 30% of the population are former Mozambicans who migrated there in the 1980s.^15^ The urban site, Jouberton Township in Klerksdorp, is part of the municipality of Matlosana in North West Province, with a population of approximately 180,000 people.

**Figure 1:**
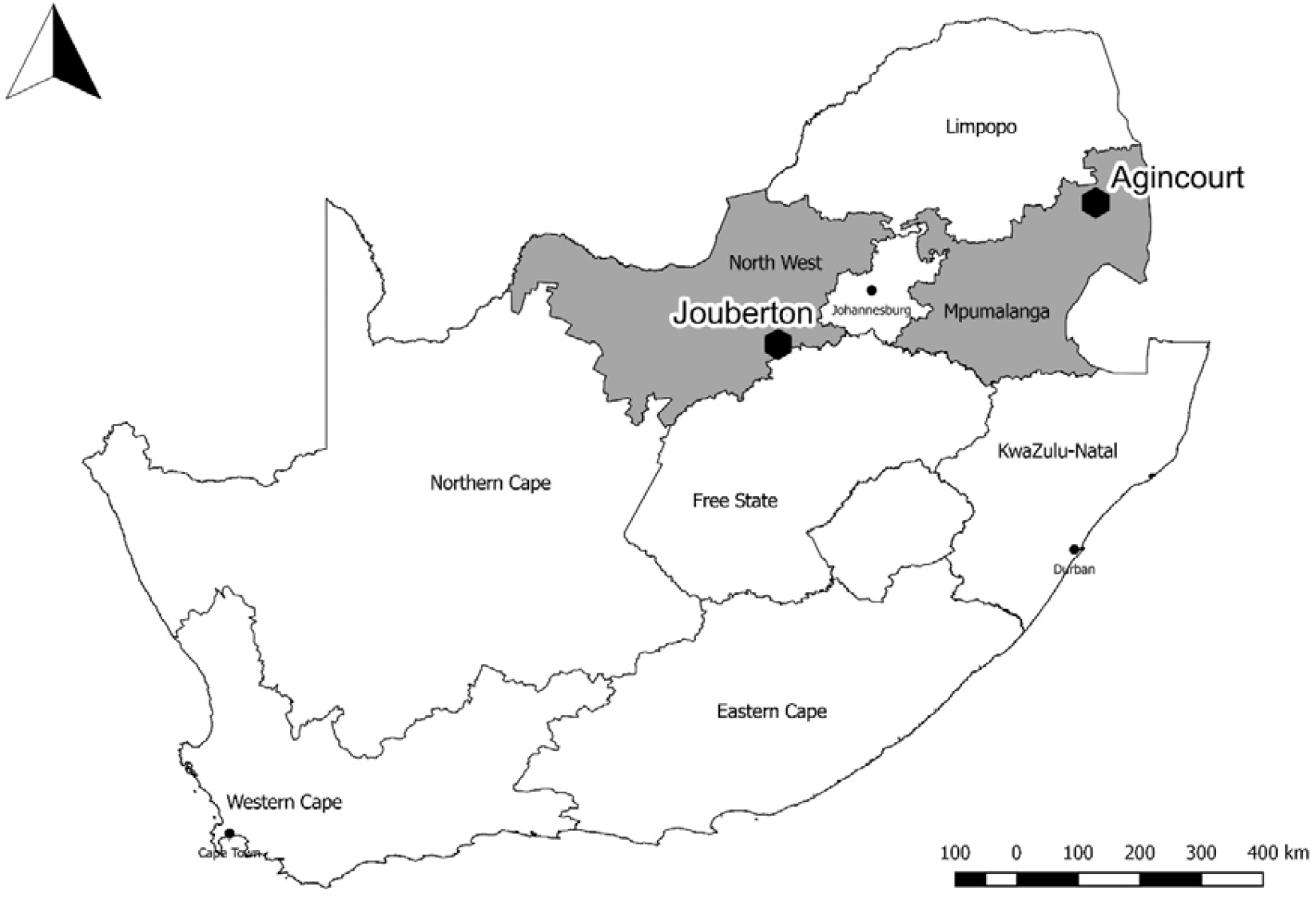
Location of rural (Agincourt) and urban (Jouberton) study sites in South Africa

We aimed to enrol approximately 1,500 individuals (approximately 500 individuals per year) over 3 consecutive influenza and RSV seasons to allow the estimation of 20% risk of infection and a 10% risk of illness with 95% confidence intervals and 5% absolute precision. Assuming an average household size of 5 individuals and a loss to follow-up of 10%, based in previous studies,^17^ we aimed to enrol approximately 55 households with >2 household members per site each year with at least 50% having at least one child aged <5 years in the house.

In rural Agincourt, each year we purposively selected two different villages within the HDSS. Within these villages, we randomly selected households with >2 members from an enumerated list obtained from the HDSS. In urban Jouberton township, we generated a list of 450 random global positioning system (GPS) coordinates located within a polygon defining the township boundaries using Google Earth. Study staff navigated to the location represented by the coordinates and selected the nearest house. If there was no dwelling within 30 meters the coordinates were discarded. Households were approached consecutively until the desired sample size was reached. If a household withdrew during January-April of each year it was replaced by a new household, selected consecutively, for the remaining follow-up period.

At each household with >2 members, study staff requested permission from the head of household to inform members about the study purpose, risks and benefits. If the head of household was a minor or unavailable after three attempts, the household was excluded. Written informed consent was required to participate in the study from all household members aged ≥18 years; assent was required from children aged 7-17 years, and consent from a parent/guardian of children aged <18 years. We included households where ≥80% of household members consented.

### Frequency of follow-up

Each year a new cohort of individuals was enrolled and following enrolment, all participants and households had a period of active twice-weekly follow up for 6-10 months. During the year of active follow up, participants received twice-weekly scheduled follow-up visits (once during Monday-Wednesday and again during Thursday-Saturday) to the household during May-October in 2016 (due to delayed start in the first study year), and January-October in 2017 and 2018 (Figure 2) for the collection of symptom data and nasopharyngeal swabs.

**Figure 2:**
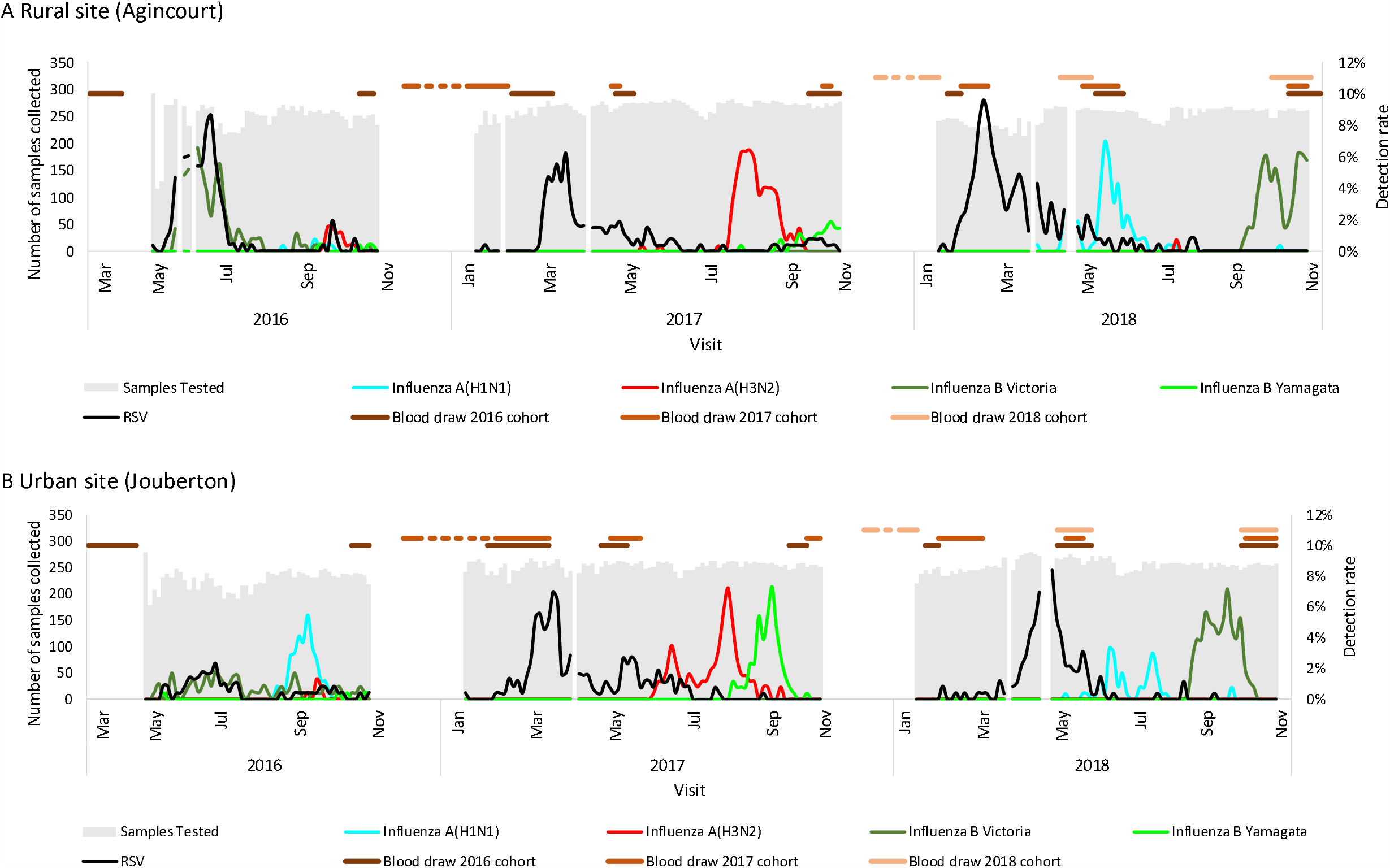
Weekly numbers of nasopharyngeal samples tested and influenza and respiratory syncytial virus (RSV) detections and timing of serology blood draws, a rural and an urban site, South Africa, 2016-2018

Household surveys were conducted once during the follow-up period to evaluate household income, housing quality, oropharyngeal carriage of meningococcus, *Corynebacterium diphtheriae* and Group A streptococcus and presence of *S. pneumoniae* DNA in blood by PCR. Serum samples were also collected at enrolment, before the influenza season, and at the end of the active follow up period. In addition, sera were also collected from the 2016 and 2017 cohorts in subsequent years (Figure 2, Supplementary Figure 1). Environmental assessments including respirable particulate matter and temperature were undertaken twice a year (summer and winter) (Table 1).

**Table 1:**
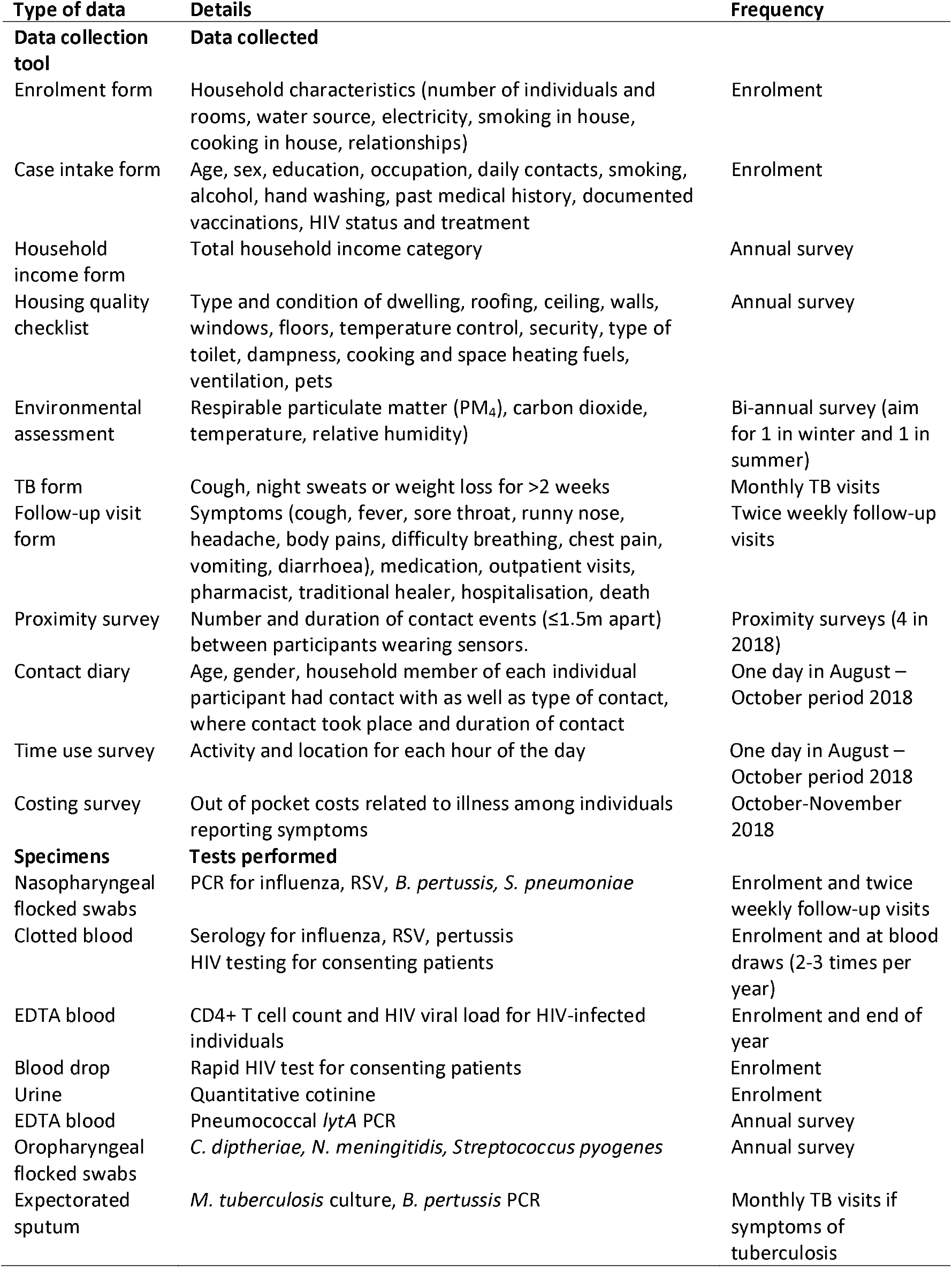
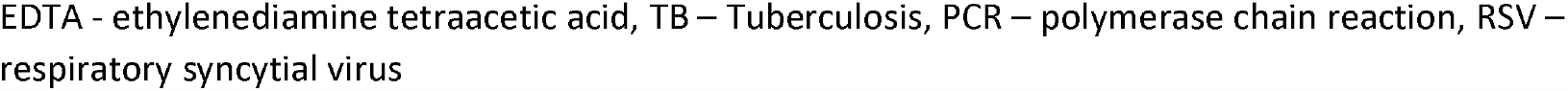
Data and specimens collected on exposures and outcomes in the PHIRST Study, South Africa, 2016-2018

### Baseline, symptom and health contact data

Data were collected using REDCap (Research Electronic Data Capture)^18^. Following enrolment, a baseline questionnaire was completed for each household including information on household members, relationships, sleeping arrangements, and housing. For each individual, we collected baseline information on demographics, underlying illnesses, vaccinations, and occupation. During the twice-weekly follow-up phase, at each visit, for each participant, a questionnaire assessing presence of symptoms, absenteeism, and health system contacts was completed and nasopharyngeal (NP) swabs were collected regardless of the presence or absence of symptoms (Table 1). Field workers were trained in the identification of respiratory signs and symptoms at the beginning of each year. Symptoms assessed at twice-weekly visits included fever (self-reported or measured tympanic temperature ≥38⍰C), cough, difficulty breathing, sore throat, nasal congestion, chest pain, muscle aches, headache, vomiting or diarrhoea. In 2017 and 2018, procedures for collection of symptom data were improved following review of symptom data from 2016. These improvements included simplification of the symptom collection form, and monthly training of fieldworkers on signs and symptoms identification and recording and frequent emphasis of the importance of symptom reporting to participants.

### Laboratory measurements

NP samples were collected using flexible nasopharyngeal nylon flocked swabs (PrimeSwab™, Longhorn Vaccines & Diagnostics, San Antonio, Texas, USA), and placed in PrimeStore® Molecular Transport Medium (MTM) (Longhorn Vaccines & Diagnostics, San Antonio, Texas, USA) and transported at 2-8°C to the National Institute for Communicable Diseases (NICD) in Johannesburg, where the samples were tested, aliquoted and stored at 2-8°C before freezing at −70°C. Nucleic acids were extracted from PrimeStore® MTM using the Roche MagNA Pure 96 instrument (Roche, Mannheim, Germany) according to the manufacturer’s instructions. NP samples were tested for RSV and influenza A and B viruses by real-time RT-PCR using the FTD Flu/RSV detection assay (Fast Track Diagnostics, Luxembourg). Influenza A positive samples were subtyped using the CDC influenza A (H1/H3/H1pdm09) subtyping kit and influenza B lineage was determined using the CDC B/Yamagata-B/Victoria lineage typing kit (available through International Reagent Resource Program; www.internationalreagentresource.org) using SuperScript™ III One-Step RT-PCR System with Platinum™ *Taq* DNA Polymerase (ThermoFisher, Waltham, Massachusetts, United States) ^19^. RSV A and B subgroups were determined by an in-house real-time RT-PCR.^20,21^ Twice-weekly NP samples were tested for *S. pneumoniae* using an in-house singleplex (*lytA*) quantitative real-time PCR assay.^22^ NP and sputum samples were tested for *Bordetella* spp. (including *B. pertussis, B. parapertussis, B. bronchiseptica* and *B. holmesii*) by a combination of a triplex and singleplex real-time PCR assays and results interpreted as previously described.^23^

Clotted blood samples from serology surveys collected in vacutainer tubes were centrifuged, aliquoted, and stored frozen before being sent in batches to NICD on dry ice. Hemagglutination inhibition (HAI) assays using turkey red blood cells were performed to determine serological reactivity titres for serum samples against influenza.^24^ Virus strains for testing were selected based on the Southern Hemisphere vaccine strains and strains predominantly circulating in South Africa during each year. Cultures from circulating strains representing the following subtypes and lineages were prepared in Madin-Darby Canine Kidney cells: A/Singapore/NFIMH-16-0019/2016-like for influenza A (H3N2), A/Michigan/45/2015-like for influenza A(H1N1)pdm09, B/Brisbane/60/2008-like for influenza B Victoria lineage and B/Phuket/3073/2013-like for influenza B Yamagata lineage. RSV antibody titres were determined using an enzyme-linked immunosorbent assay (ELISA) (EUROIMMUN Anti-RSV IgG; Lübeck, Germany).^25^ Serology for pertussis antibody was performed using the commercially available anti-Pertussis toxin ELISA (EUROIMMUN, Lübeck, Germany).^26^

Urine cotinine tests were performed using the IMMULITE® 1000 Nicotine Metabolite Assay Kit (Siemens Medical Solutions Diagnostics, Gly Rhonwy, UK). Sputum samples were collected at baseline from individuals who could expectorate and thereafter from any individual with fever, cough, or weight loss for >2 weeks, assessed monthly. Sputum samples were tested for tuberculosis using GeneXpert MTB/RIF (Cepheid, Sunnyvale, CA, USA) and cultured using the Mycobacteria Growth Indicator Tube (MGIT) 960 instrument (Becton Dickinson, East Rutherford, NJ, USA).

Tuberculin skin tests were performed twice annually 6-10 months apart. Five units of purified tuberculin was injected intradermally on the volar aspect of the forearm. Results were assessed 48-72 hours thereafter, for induration and, if present, study staff measured the diameter in millimeters transverse to the axis of the forearm. HIV testing, according to the double rapid test algorithm in place in each province was offered to all participants older than 10 years of age.^27^ Participants were considered HIV-infected if they had one of the following during the follow up period: two positive rapid HIV tests, evidence of a positive HIV laboratory result or evidence of antiretroviral treatment. Participants were considered HIV-uninfected if they had a documented negative HIV test result during the study. A documented HIV negative status for the mother was considered confirmation of HIV negative status for a child aged <10 years. Infants were defined as HIV-exposed but uninfected if they were HIV-uninfected, but the mother was HIV-infected. HIV-infection was confirmed by PCR in children aged <18 months. For all HIV-infected individuals, specimens were collected for CD4+ T cell and quantitative HIV viral load testing. Patients newly diagnosed with HIV were referred to the local clinic for assessment and initiation of antiretroviral treatment.

### Environmental assessment

Environmental monitoring was conducted in a convenience subset of 150 households for one week each year during summer and winter. Respirable particulate matter <4µm diameter (PM4) concentrations were measured indoors using a stationary photometric monitor (DustTrak II Model 8530, TSI Incorporated, Shoreview, MN, USA) and gravimetric filter sampling to measure the quantity of airborne particulate matter. During the same period, one member of each household carried a personal exposure monitoring device (SidePak AM510, TSI Incorporated, Shoreview, MN, USA) throughout the daytime. Indoor carbon-dioxide and ambient PM4 (ES-642, MetOne Instruments, Inc, Grants Pass, OR, USA) levels were measured during 2018. Thermochron® iButton sensors (Maxim Integrated, San Jose, California, USA) were located in the indoor (all households) and ambient (subset) environments to measure temperature (Model DS1921G-F5) and relative humidity (Model RS1923l-F5 in 2018) concurrently with the air pollution monitoring in 2016 and continuously during 2017-2018.

### Housing quality survey

Information on housing type, construction, materials and condition, water sources, water security and water storage, fuel use and expenditure for cooking, space and water heating, waste removal services, visible dampness and smoking practices was collected annually for all households.

### Proximity and contact study

In 2018, four surveys of household contact using proximity monitors (www.sociopatterns.org) were conducted to capture information on intra-household contact patterns for three seasons (summer, autumn, and winter). To measure contacts of participants of the study outside the home, participants were interviewed by field workers to complete a contact diary and time-use questionnaire for one day between August and October 2018.

### Costing survey

We surveyed all symptomatic household members during August-October 2018 to assess cost of medically attended and non-medically attended illness episodes.

## Findings to date

### Household and individual characteristics

From 2016 through 2018 at both sites combined, 881 households were approached: for 409 (46%) households, the head of household agreed to participate and 327 (78%) of these were included in the final analysis (Figure 3a and 2b). There were 1,861 individuals residing in the 327 included households, of whom, 1,684 (90%) consented and were included in the final analysis. Reasons for non-inclusion are shown in Figure 3a and b. A higher percentage of approached houses were included at the rural site (159/267, 60%) compared to the urban site (168/614, 27%). This is because rural site houses with >2 members were pre-selected from the HDSS database and a higher proportion of household members consented to participate in the study among eligible households (209/252, 83% vs 200/326, 61%, p<0.001).

**Figure 3:**
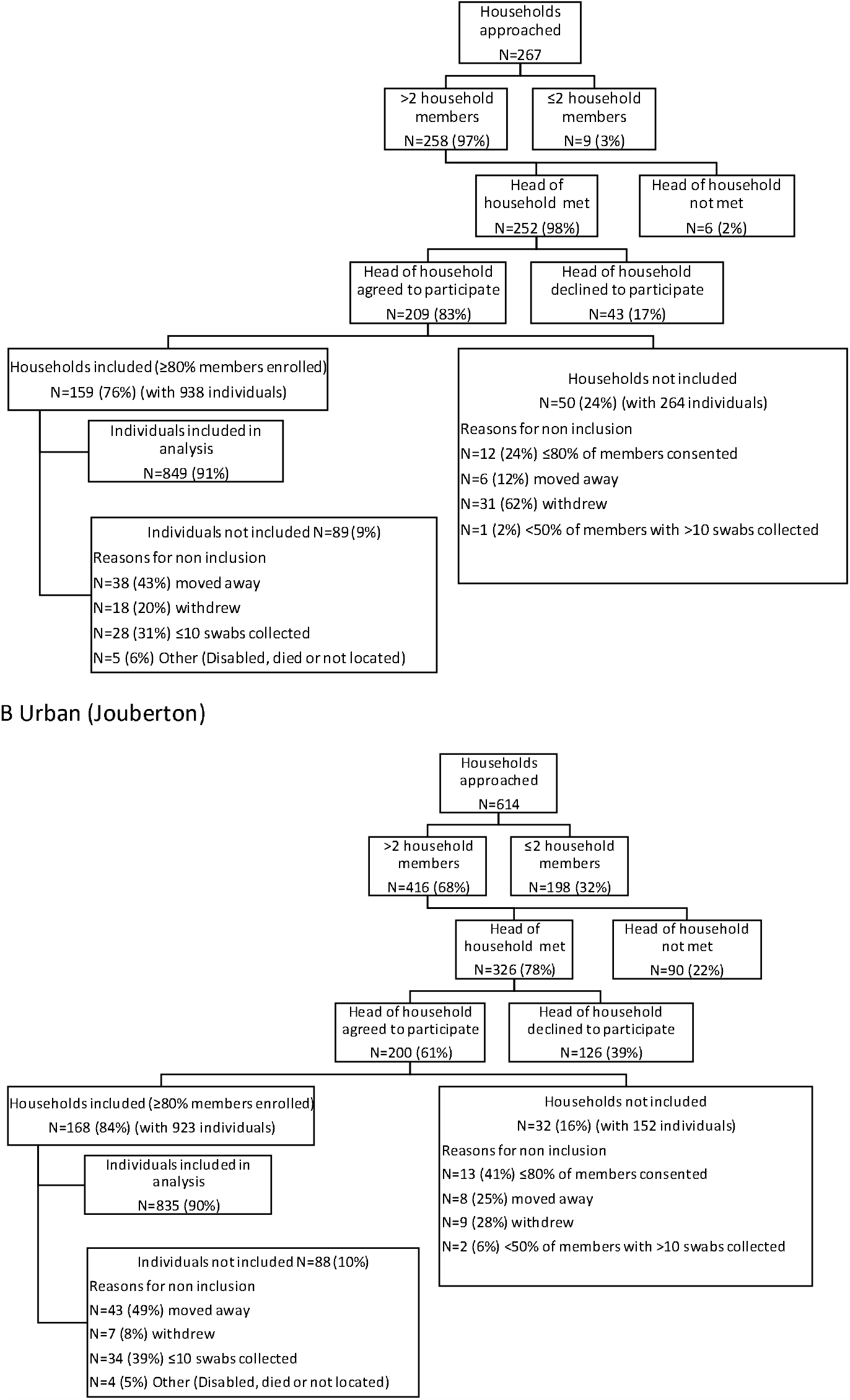
Flow chart of participant enrolment at the rural and urban sites, South Africa, 2016-2018 A Rural (Agincourt) Individuals or households were not included if they withdrew or moved away before completing >10 visits with collection of a nasopharyngeal swab Bars indicate timing of serology blood draws. Grey bars indicate number of nasopharyngeal swabs tested each visit. Blood draw dates in year **2016** were: draw 1 (16 March – 15 April), draw 2 (17 October – 24 November). Blood draw dates in year **2017** were: 2016 cohort draw 1 (6 February – 4 March), draw 2 (3 May – 26 May), draw 3 (2 October – 18 October); 2017 cohort draw 1 (24 November 2016 to 24 February 2017), draw 2 (2 May – 26 May) draw 3 (12 October – 27 October). Blood draw dates in year **2018** were: 2016 cohort draw 1 (29 January – 9 February), draw 2 (4 May – 6 June), draw 3 (1 October to 5 November); 2017 cohort draw 1 (12 February – 9 March), draw 2 (4 May – 4 June), draw 3 (17 October – 31 October); 2018 cohort draw 1 (28 November 2017 – 24 January 2018), draw 2 (21 April – 1 June 2018), draw 3 (1 October – 31 October).

At the rural site, characteristics of included households were similar to those of households within the HDSS that were not included (Table 2a). However, when compared to individuals from the HDSS who were not included, included individuals were less likely to be aged 15-44 years, male, employed, or have completed secondary education. This likely reflects migrant worker patterns and the fact that within included households, males and individuals aged 15-44 years were less likely to participate in the study (Supplementary table 2). At the urban site, characteristics of households and individuals included in PHIRST were compared to those of residents of Jouberton Township from the 2011 census^28^ (Table 2b). Compared to the general population, included households were more likely to be formal (brick houses on a municipal stand) houses rather than shacks within informal settlements, possibly because larger stands are more likely to be sampled using GPS coordinates. HIV prevalence among included individuals aged 15-49 years with available data was 27% (99/370) at the rural site and 29% (97/329) at the urban site, compared to 23% (95% confidence interval (CI) 20-26%) and 23% (95% CI 18-28) reported for Mpumalanga (rural site) and North West (urban site) Provinces in 2017, respectively.^29^

**Table 2a:** Characteristics of included participants and households in PHIRST during 2016-2018 at the rural site (Agincourt) compared to those not included, using data from the 2017 Agincourt health and socio-demographic surveillance system site (HDSS) census

| Characteristic | Included<br>n/N (%) or number<br>(range) | Not included<br>n/N (%) or number<br>(range) | p |
| --- | --- | --- | --- |
| **Household level** |  |  |  |
| Mean number (range) members in household | 4 (1-20) | 4 (1-15) | 0.479 |
| Modern house (vs traditional) | 136/136 (100) | 16,738/16,785(99.7) | 0.587 |
| Toilet site |  |  |  |
| In house | 2/136 (1) | 268/16,782 (2) | 0.658 |
| In yard | 121/136 (89) | 15,249/16,782 (91) |  |
| Other | 13/136 (10) | 1,265/16,782 (8) |  |
| Type of toilet |  |  |  |
| Modern | 3/136 (2) | 245/16,780 (1) | 0.151 |
| VIP | 7/136 (5) | 1,841/16,780 (11) |  |
| Pit latrine | 116/136 (85) | 13,676/16,780 (82) |  |
| Other | 10/136 (7) | 1,018/16,780 (6) |  |
| Source of energy for cooking |  |  |  |
| Electricity | 58/136 (43) | 8,929/16,781 (53) | 0.050 |
| Wood | 77/136 (57) | 7,759/16,781 (46) |  |
| Gas/Paraffin (kerosene)/Other | 1/136 (1) | 93/16,781 (1) |  |
| **Individual level** |  |  |  |
| Age group (years) |  |  |  |
| <1 | 2/791 (0.3) | 1,302/99,645 (1) | <0.001 |
| 1-4 | 114/791 (14) | 9,079/99,645 (9) |  |
| 5-14 | 296/791 (37) | 22,568/99,645 (23) |  |
| 15-44 | 274/791 (35) | 51,128/99,645 (51) |  |
| 45-64 | 70/791 (9) | 9,966/99,645 (10) |  |
| 65+ | 35/791 (4) | 4,602/99,645 (5) |  |
| Female sex | 499/791 (63) | 52,972/98,640 (54) | <0.001 |
| Level of education<sup>b</sup> |  |  |  |
| None | 173/704 (25) | 17,500/88,149 (20) | <0.001 |
| Primary | 265/704 (38) | 24,238/88,149 (27) |  |
| Some secondary | 202/704 (29) | 26,088/88,149 (30) |  |
| Completed secondary | 64/704 (9) | 20,322/88,149 (23) |  |
| Post-secondary | 0/704 (0) | 1/88,149 (0) |  |
| Currently working<sup>c</sup> | 60/495 (12) | 19,267/74,923 (26) | <0.001 |

VIP – Ventilated improved pit latrine <sup>a</sup>Only households and individuals with data available in the Agincourt HDSS included, for individual level analysis only permanent (non-migrant) household members included. Data are mean (range) or n/N (%) <sup>b</sup>The highest level an individual has attained at the time of observation <sup>c</sup>Individuals aged ≥17 years

**Table 2b:** Characteristics of included participants at the urban site (Jouberton) during 2016-2018 from PHIRST database and characteristics of the general population from the 2011 Census^28^

| Characteristic | Enrolled<br>n/N (%) or<br>number (range) | General population <sup>a</sup><br>n/N (%) or number (range) |
| --- | --- | --- |
| **Household level** |  |  |
| Mean number (range) members in household | 5 (3-14) | 4 (3-5)<sup>d</sup> |
| Mean number (range) rooms in household | 5 (2-11) | 4 (1-18) |
| Monthly household income |  |  |
| ≤R800 (<\$54) | 18/167 (11) | 7,119/32,136 (22) |
| R801-R1600 (\$55-\$108) | 35/167 (21) | 2,835/32,136 (9) |
| R1601-R3200 (\$109-\$116) | 49/167 (29) | 6,507/32,136 (20) |
| R3201-R6400 (\$117-\$232) | 34/167 (20) | 7,251/32,136 (23) |
| R6401-R12800 (\$233-\$464) | 9/167 (5) | 4,851/32,136 (15) |
| >R12800 (>\$464) | 5/167 (3) | 3,558/32,136 (11) |
| Did not disclose | 17/167 (10) | 15/32,136 (0) |
| Type of dwelling |  |  |
| Formal house | 158/167 (95) | 23,379/32,130 (73) |
| Informal dwelling | 6/167 (4) | 6,777/32,130 (21) |
| Formal dwelling in backyard | 0/167 (0) | 882/32,130 (3) |
| Flat | 0/167 (0) | 225/32,130 (1) |
| Traditional dwelling | 0/167 (0) | 93/32,130 (0) |
| Other | 3/167 (2) | 774/32,130 (2) |
| **Individual level** |  |  |
| Age group (years) |  |  |
| <1 | 27/858 (3) | 2,664/111,936 (2) |
| 1-4 | 89/858 (10) | 10,590/111,936 (9) |
| 5-14 | 238/858 (28) | 21,051/111,936 (19) |
| 15-44 | 334/858 (39) | 54,297/111,936 (49) |
| 45-64 | 124/858 (14) | 18,684/111,936 (17) |
| 65+ | 46/858 (5) | 4,650/111,936 (4) |
| Female sex | 487/859 (57) | 57,129/111,939 (51) |
| Level of education<sup>b</sup> |  |  |
| No schooling | 26/443 (6) | 7,872/98,442 (8) |
| Primary schooling | 116/443 (26) | 34,767/98,442 (35) |
| Some secondary | 207/443 (47) | 35,295/98,442 (36) |
| Secondary completed | 81/443 (18) | 18,015/98,442 (18) |
| Post-secondary | 13/443 (3) | 2,493/98,442 (3) |
| Currently working<sup>c</sup> | 154/444 (35) | 24,642/72,981 (34) |

<sup>a</sup>Data on general population obtained from census 2011<sup>28</sup> unless otherwise indicated. Data are median (IQR) or n/N (%) <sup>b</sup>>18 years PHIRST, >5 years in census, <sup>c</sup>>18 years PHIRST, >15 years in census, <sup>d</sup>Source <sup>17</sup>

Among 327 households included in the study, the median household size was 5 individuals (interquartile range 3-10) and 160 (49%) reported crowding (>2 people/sleeping room) (Table 3). Among 1,684 study participants, 16% were aged <5 years, 32% aged 5-14 years and 60% were female. Compared to the urban site, households in the rural site were more likely to have a child aged <5 years in the house and to use wood as fuel for cooking, and less likely to report smoking in the house or to have a handwashing place with water. Compared to the urban site, individuals in the rural site were more likely to be aged <15 years, female, unemployed, and less likely to drink alcohol or smoke.

**Table 3:** Baseline characteristics of households and participants included in the final cohort by site, a rural and an urban site in South Africa, PHIRST Study, 2016-2018

| Characteristic | Overall<br>n (%) or<br>median<br>(IQR) | Rural<br>n (%) or<br>median<br>(IQR) | Urban<br>n (%) or<br>median<br>(IQR) | OR (95% CI) | p |
| --- | --- | --- | --- | --- | --- |
| **Household level characteristics** | **N=327** | **N=159** | **N=168** |  |  |
| Year |  |  |  |  |  |
| 2016 | 100 (31) | 50 (31) | 50 (30) | Reference | 0.932 |
| 2017 | 109 (33) | 53 (33) | 56 (33) | 1.1 (0.6-1.8) |  |
| 2018 | 118 (36) | 56 (35) | 62 (37) | 1.1 (0.6-1.9) |  |
| Number of household members |  |  |  |  |  |
| 3-5 | 196 (60) | 89 (56) | 107 (64) | Reference | 0.341 |
| 6-10 | 119 (36) | 63 (40) | 56 (33) | 0.7 (0.5-1.2) |  |
| >10 | 12 (4) | 7 (4) | 5 (3) | 0.6 (0.2-1.9) |  |
| Number of household members | 5 (3-10) | 5 (3-11) | 5 (3-10) | Not estimated | 0.442 |
| Number of rooms | 5 (2-9) | 5 (1-10) | 5 (2-8) | Not estimated | 0.453 |
| Number of rooms for sleeping | 2 (1-4) | 3 (1-4) | 2 (1-4) | Not estimated | 0.256 |
| Crowding (>2 people/sleeping room) | 160 (49) | 83 (52) | 77 (46) | 0.8 (0.5-1.2) | 0.250 |
| Child aged <5 years in house | 225 (69) | 141 (89) | 84 (50) | 0.1 (0.1-0.2) | <0.001 |
| HIV-infected household member | 172 (53) | 80 (50) | 92 (55) | 1.2 (0.8-1.8) | 0.421 |
| Cigarette smoke in house | 71 (22) | 18 (11) | 53 (32) | 3.6 (2.0-6.5) | <0.001 |
| Main water source tap inside (vs tap outside) | 154 (47) | 71 (45) | 83 (49) | 0.8 (0.5-1.3) | 0.390 |
| Handwashing place with water in house | 264 (81) | 100 (63) | 164 (98) | 24.2 (8.6-68.3) | <0.001 |
| Main fuel for cooking |  |  |  |  |  |
| Electricity | 244 (75) | 89 (56) | 155 (93) | 121.9 (16.6-892.8) | <0.001 |
| Wood | 71 (22) | 70 (44) | 1 (1) | Reference |  |
| Paraffin(kerosene)/gas/other | 10 (3) | 0 (0) | 10 (6) | Not estimated |  |
| Monthly household income<sup>a</sup> |  |  |  |  |  |
| ≤R800 (<\$54) | 39 (12) | 21 (14) | 18 (11) | Reference | 0.807 |
| R801-R1600 (\$55-\$108) | 75 (24) | 36 (23) | 39 (24) | 1.3 (0.6-2.7) | |
| R1601-R3200 (\$109-\$116) | 109 (34) | 56 (36) | 53 (33) | 1.1 (0.5-2.3) | |
| R3201-R6400 (\$117-\$232) | 70 (22) | 32 (21) | 38 (23) | 1.4 (0.6-3.0) | |
| R6401-R12800 (\$233-\$464) | 18 (6) | 8 (5) | 10 (6) | 1.5 (0.5-4.5) | |
| >R12800 (>\$464) | 7 (2) | 2 (1) | 5 (3) | 0.9 (0.5-16.8) | |
| **Individual level characteristics** | **N=1684** | **N=849** | **N=835** |  |  |
| Age group (years) |  |  |  |  |  |
| <1 | 36 (2) | 15 (2) | 21 (3) | 2.5 (1.2-5.1) | <0.001 |
| 1-4 | 243 (14) | 156 (18) | 87 (10) | Reference |  |
| 5-14 | 547 (32) | 309 (36) | 238 (29) | 1.4 (1.1-1.9) |  |
| 15-44 | 590 (35) | 265 (31) | 325 (39) | 2.1 (1.6-3.0) |  |
| 45-64 | 195 (12) | 74 (9) | 121 (14) | 2.9 (2.0-4.3) |  |
| 65+ | 73 (4) | 30 (4) | 43 (5) | 2.6 (1.5-4.4) |  |
| Female sex | 1009 (60) | 533 (63) | 476 (57) | 0.8 (0.6-0.9) | 0.016 |
| Year |  |  |  |  |  |
| 2016 | 542 (32) | 280 (33) | 262 (31) | Reference | 0.768 |
| 2017 | 577 (34) | 289 (34) | 288 (34) | 1.1 (0.8-1.3) |  |
| 2018 | 565 (34) | 280 (33) | 285 (34) | 1.1 (0.9-1.4) |  |
| Level of education<sup>b</sup> |  |  |  |  |  |
| No schooling | 90 (12) | 65 (22) | 25 (6) | Reference | <0.001 |
| Primary schooling | 158 (22) | 60 (20) | 98 (23) | 4.2 (2.4-7.5) |  |
| Some secondary | 285 (39) | 85 (28) | 200 (47) | 6.1 (3.6-10.3) |  |
| Secondary completed | 174 (24) | 87 (29) | 87 (21) | 2.6 (1.5-4.5) |  |
| Post-secondary | 19 (3) | 5 (2) | 14 (3) | 7.3 (2.4-22.3) |  |
| Employment<sup>b</sup> |  |  |  |  |  |
| Unemployed | 400 (55) | 182 (60) | 218 (51) | Reference | 0.014 |
| Employed | 267 (37) | 88 (29) | 179 (42) | 1.7 (1.2-2.3) |  |
| Student | 59 (8) | 32 (11) | 27 (6) | 0.7 (0.9-1.5) |  |
| Reported alcohol use<sup>c</sup> | 337 (39) | 57 (15) | 280 (57) | 7.3 (5.2-10.2) | <0.001 |
| Reported current cigarette smoking<sup>c</sup> | 139 (16) | 22 (6) | 117 (24) | 4.9 (3.0-7.9) | <0.001 |
| Reported current snuff smoking<sup>c</sup> | 97 (11) | 5 (1) | 92 (19) | 17.0 (6.8-42.1) | <0.001 |
| Reported current any smoking<sup>c</sup> | 242 (28) | 28 (8) | 214 (44) | 9.5 (6.2-14.4) | <0.001 |
| Cigarette smoke inside house<sup>d</sup> | 99 (41) | 7 (25) | 92 (43) | 2.3 (0.9-5.5) | 0.048 |
| Urine cotinine (all ages)<sup>e</sup> |  |  |  |  |  |
| Negative | 649 (42) | 530 (68) | 119 (16) | Reference | <0.001 |
| Passive exposure | 660 (43) | 222 (28) | 438 (57) | 8.8 (6.8-11.4) |  |
| Active smoking | 241 (16) | 31 (4) | 210 (27) | 30.2 (19.7-46.2) |  |
| Unknown | 134 | 66 | 68 | Not included |  |
| HIV status<sup>e</sup> |  |  |  |  |  |
| Uninfected | 1379 (85) | 715 (86) | 664 (83) | Reference | 0.158 |
| Infected | 249 (15) | 117 (14) | 132 (17) | 1.2 (0.9-1.6) |  |
| Unknown | 56 | 17 | 39 | Not included |  |
| Previous tuberculosis | 88 (5) | 15 (2) | 73 (9) | 5.3 (3.0-9.4) | <0.001 |
| Current tuberculosis | 24 (1) | 3 (<1) | 21 (3) | 7.3 (2.2-24.5) | <0.001 |
| Other underlying illness<sup>f</sup> | 50 (3) | 5 (1) | 45 (5) | 9.6 (3.8-24.3) | <0.001 |
| Influenza vaccination current year | 2 (<1) | 1 (<1) | 1 (<1) | 1.0 (0.1-16.2) | 0.990 |
| Pneumococcal vaccine up to date for age<sup>g</sup> |  |  |  |  |  |
| Yes | 221 (81) | 137 (81) | 84 (80) | 0.2 (0.1-1.3) | 0.182 |
| No | 7 (3) | 2 (1) | 5 (5) | Reference |  |
| No data | 46 (17) | 30 (18) | 16 (15) | 0.2 (0.1-1.2) |  |
| DTaP-IPV/Hib vaccine up to date for age<sup>g</sup> |  |  |  |  |  |
| Yes | 222 (81) | 135 (80) | 87 (83) | 0.9 (0.2-3.9) | 0.735 |
| No | 7 (3) | 4 (2) | 3 (3) | Reference |  |
| No data | 45 (16) | 30 (18) | 15 (14) | 0.7 (0.1-3.4) |  |

<sup>a</sup> Data available for 318 households, 155 rural and 163 urban <sup>b</sup> Individuals aged ≥18 years N=726, 302 at rural and 424 at urban site <sup>c</sup> Self-reported, individuals aged ≥15 years N=858, 369 at rural and 489
at urban site <sup>d</sup>Amongst those reporting any current smoking <sup>e</sup>% and p value among individuals with known status <sup>f</sup>Self-reported history of asthma, lung disease, heart disease, stroke, spinal cord injury, epilepsy, organ transplant, immunosuppressive therapy, organ transplantation, cancer, liver disease, renal disease or diabetes <sup>g</sup>Individuals aged <5 years N=274, 169 at rural site and 105 at urban site, 229 with available vaccination data, 139 at the rural site and 90 at the urban site

Among 1,684 included individuals, 1,605 (95%) were present at end of the twice weekly follow-up phase. Of 79 lost to follow-up, 53 (67%) left the study sites, 21 (27%) withdrew and 5 (6%) died (Table 4). Just over half of the individuals in the 2016 and 2017 cohorts completed 3 serology blood draws in the years after completion of the swabbing phase. Individuals lost to follow-up were more likely to be aged 15-44 years, possibly due to economic migration, compared to those completing follow-up (Supplementary table 3).

**Table 4:** Follow-up rates by site and year among 1684 individuals included in the analysis of the PHIRST Study, South Africa, 2016-2018

| Characteristic | Site |  |  | Year |  |  |
| --- | --- | --- | --- | --- | --- | --- |
|  | Overall<br>N (%) | Rural<br>N (%) | Urban<br>N (%) | 2016<br>N (%) | 2017<br>N (%) | 2018<br>N (%) |
| **Twice-weekly follow-up phase** |  |  |  |  |  |  |
| **Individuals** |  |  |  |  |  |  |
| Total included in analysis | 1,684 | 849 | 835 | 542 | 577 | 565 |
| Initially enrolled<sup>a</sup> | 1,555<br>(92) | 790 (93) | 765 (92) | 505 (93) | 551 (95) | 499 (88) |
| Late enrolments for replacement or in-migration to household | 129 (8) | 59 (7) | 70 (8) | 37 (7) | 26 (5) | 66 (12) |
| Lost to follow-up | 79 (5) | 32 (4) | 47 (6) | 21 (4) | 31 (5) | 27 (5) |
| Withdrawal n(%) | 21 (27) | 13 (41) | 8 (17) | 9 (43) | 6 (19) | 6 (22) |
| Left study site n(%) | 53 (67) | 17 (53) | 36 (77) | 12 (57) | 21 (68) | 20 (74) |
| Death n(%) | 5 (6) | 2 (6) | 3 (6) | 0 (0) | 4 (13) | 1 (4) |
| Completed follow-up | 1,605<br>(95) | 817 (96) | 788 (94) | 521 (96) | 546 (95) | 538 (95) |
| **Households** |  |  |  |  |  |  |
| Total included | 327 | 159 | 168 | 100 | 109 | 118 |
| Initially enrolled<sup>a</sup> | 308 (94) | 148 (93) | 160 (95) | 96 (96) | 105 (96) | 107 (91) |
| Late enrolments for replacement or in-migration to household | 19 (6) | 11 (7) | 8 (5) | 4 (4) | 4 (4) | 11 (9) |
| Lost to follow-up | 6 (2) | 1 (1) | 5 (3) | 1 (1) | 2 (2) | 3 (3) |
| Withdrawal n(%) | 3 (1) | 1 (1) | 2 (1) | 1 (1) | 2 (2) | 0 (0) |
| Left study site n(%) | 3 (1) | 0 (0) | 3 (2) | 0 (0) | 0 (0) | 3 (3) |
| Completed follow-up | 321 (98) | 158 (99) | 163 (97) | 99 (99) | 107 (98) | 115 (97) |
| **Serology phase individuals<sup>b</sup>** |  |  |  |  |  |  |
|  | **n/N (%)** | **n/N (%)** | **n/N (%)** |  |  |  |
| 2016 cohort: 3 blood draws in serology phase 2017 | 297/542<br>(55) | 116/280<br>(41) | 181/262<br>(69) | NA | NA | NA |
| 2016 cohort: 3 blood draws in serology phase 2018 | 295/542<br>(54) | 117/280<br>(42) | 178/262<br>(68) | NA | NA | NA |
| 2017 cohort: 3 blood draws in serology phase 2018 | 303/577<br>(53) | 118/289<br>(41) | 185/288<br>(64) | NA | NA | NA |

<sup>a</sup>Before 31 March for 2016, before 31 January for 2017 and 2018.
<sup>b</sup>Timing of blood draws indicated graphically in Figure 3. Participants in the 2016 cohort had two blood draws during 2016 and 3 each in 2017 and 2018, participants in the 2017 cohort had 3 blood draws in 2017 and 3 in 2018, participants in the 2018 cohort had 3 blood draws in 2018.
NA – Not applicable

### Samples collected and symptoms

There were 122,113 potential individual follow-up visits over the 3 years, and participants were interviewed for 105,783 (87%) of these. In 2017-2018, there were 94,786 potential visits, of which participants were interviewed for 81,943 (86%) and 81,928 (>99%) had available data on symptoms. At least one symptom was reported for 8% (6,692) of visits overall. At least one symptom over the follow-up period in 2017-2018 was reported by 89% (1,012/1,142) of individuals and was more commonly reported among children aged <5 years (97%, 180/185) compared to older individuals (5-18 years 87%, 404/466; 19-65 years 87%, 390/450; >65 years 93%, 38/41, p<0.001 Chi squared test). The commonest symptoms reported were cough (76%, 863/1,142) and runny nose (74%,841/1,142). The rate of clinic visits in 2017-2018 for acute complaints was 1.3 per 100 person weeks of follow-up (n=520) (2.1 in <5 years, 0.9 in 5-18 years, 1.3 in 19-65 years, 2.0 in >65 years) and of hospitalisations was 0.05 per 100 person weeks of follow-up (n=36)(0.09 in <5 years, 0.05 in 5-18 years, 0.10 in 19-65 years, 0.3 in >65 years).

From May 2016-December 2018 a total of 105,683 nasopharyngeal swabs, 4,217 clotted blood samples, 1,442 whole blood samples, 1,567 urine samples and 741 sputum samples were collected. Out of 105,683 nasopharyngeal swabs from follow-up visits collected and tested from 1,684 participants, in 327 households, 1,258 (1%), 1,026 (1%), 273 (<1%), 38,829 (37%) tested positive on PCR for influenza viruses, respiratory syncytial virus, pertussis and pneumococcus respectively.

### Future plans

Households enrolled in the PHIRST study during 2016-2018 were eligible for inclusion in a study of SARS-CoV-2 transmission initiated in July 2020. This study uses similar testing frequency and household selection methods to assess the community burden of SARS-CoV-2 infection and the role of asymptomatic infection in virus transmission.

## Strengths and Limitations

### Strengths

PHIRST was conducted in urban and rural African settings. Situation in a high HIV prevalence setting with high study uptake of HIV testing (97%) allows assessment of the effect of HIV on community burden and transmission dynamics of respiratory pathogens. Households were selected randomly to provide a representative sample of the community. Sampling from individuals irrespective of symptoms allows estimation of community burden, household secondary infection risk, and serial interval including asymptomatic or paucisymptomatic episodes.^30^ It also allows the estimation of the proportion of transmission from asymptomatic individuals. PHIRST utilised multiple laboratory-confirmed infection endpoints including PCR and serology which provide additional data on the community burden of these pathogens and allows evaluation of the correlation between pathogen detection and serological response in individuals of different age and HIV-infection status. Laboratory-confirmation of multiple respiratory pathogens simultaneously allows study of the effect of respiratory co-infections on disease severity and transmissibility and the interaction between different pathogens. Our twice-weekly sampling strategy was unlikely to miss many episodes of infection and allowed accurate ascertainment of first and subsequent infections from the same or different pathogens in the household. Our long period of active follow-up for each cohort allowed us to describe burden and transmission of infection within described seasons.

### Limitations

This study was not powered to assess severe outcomes (i.e. hospitalisation and death). Repeated assessment of symptoms at twice-weekly visits over an extended period may lead to possible fatigue and under-reporting by participants. High rates of migration and movement in communities under study affected follow-up rates. The study was intensive and nasopharyngeal swabs are uncomfortable for participants which may have resulted in fewer participants consenting and reduced follow-up rates. Quality of specimen collection may have varied; however, >99% of samples tested positive for the presence of human DNA. We only examined four pathogens, but other micro-organisms may be important. Samples have been stored which could allow us to implement broader multi-pathogen testing in the future. We did not test staff members for the presence of organisms under study, because a previous similar study in which staff were sampled weekly did not identify any influenza or RSV infections among field staff.^31^

## Collaborations and data statement

Primary study results for influenza as well as a description of the quality of housing at the two sites have been prepared and submitted to international peer-reviewed journals. Analysis of the data for other pathogens is planned to be completed by December 2021. Additional modelling and serologic studies will be concluded within 3 years and primary de-identified data should be publicly available no later than November 2023.

The investigators welcome enquiries about possible collaborations and access to the data set. Investigators interested in more details about this study, or in accessing these resources, should contact the principle investigator, Prof Cheryl Cohen, at NICD.

## Data Availability

Primary study results for influenza as well as a description of the quality of housing at the two sites have been prepared and submitted to international peer-reviewed journals. Analysis of the data for other pathogens is planned to be completed by December 2021. Additional modelling and serologic studies will be concluded within 3 years and primary de-identified data should be publicly available no later than November 2023.
The investigators welcome enquiries about possible collaborations and access to the data set. Investigators interested in more details about this study, or in accessing these resources, should contact the principle investigator, Prof Cheryl Cohen, at NICD.

## Further details

### Ethical review

The protocol was registered on clinicaltrials.gov on 6 August 2015 (Reference NCT02519803) and was approved by the University of the Witwatersrand Human Research Ethics Committee (Reference 150808) and the U.S. CDC’s Institutional Review Board relied on the local review (#6840). Participants provided individual written consent or assent prior to enrolment and received a grocery store voucher of ZAR25-30 (USD 2-2.5) per visit for their time.

### Patient and public involvement

Both study sites have community advisory boards (CAB) consisting of representatives from community-based and faith-based organizations who were involved in the planning of the PHIRST study. The CABs meet regularly and give advice on protocols, consents and recruitment plans and also provide feedback to communities on results of studies. In addition, feedback sessions on study findings were held for participating families.

### Funding

The study was funded through a cooperative agreement with the United States Centers for Disease Control and Prevention (CDC) (grant number 1U01IP001048). Testing for RSV and pneumococcus was supported by the Bill and Melinda Gates Foundation (Grant number: OPP1164778). Testing for *B. pertussis* was supported by Sanofi Pasteur (Grant number: PER00059). The Agincourt Health and Socio-Demographic Surveillance System is a node of the South African Population Research Infrastructure Network (SAPRIN) and is supported by the National Department of Science and Innovation, the Medical Research Council and the University of the Witwatersrand, South Africa, and the Wellcome Trust, UK (grants 058893/Z/99/A; 069683/Z/02/Z; 085477/Z/08/Z; 085477/B/08/Z).

## Acknowledgements

We would like to acknowledge the study participants as well as field and laboratory staff who worked tirelessly to make the study a success.

## Author Statement

CC, MM, TM, JM, FKT, OH, NW, NAM, KK, AvG, ST: Contributed to conception or design of protocol

All co-authors: Contributed to acquisition, analysis or interpretation of data for the work

FKT, OH, NW, MC, AB, LM, AvG: Laboratory testing of samples LL, MM, KM, FW, SN: Enrolment and follow-up of participants

CC, MM, ST: Drafting the work or revising it critically for important intellectual content

All co-authors: Final approval of the version to be published

All co-authors: Agreement to be accountable for all aspects of the work in ensuring that questions related to the accuracy or integrity of any part of the work are appropriately investigated and resolved

## Disclaimer

The findings and conclusions in this report are those of the author(s) and do not necessarily represent the official position of the funding agencies.

## Supplementary tables and figures

**Supplementary table 1:**
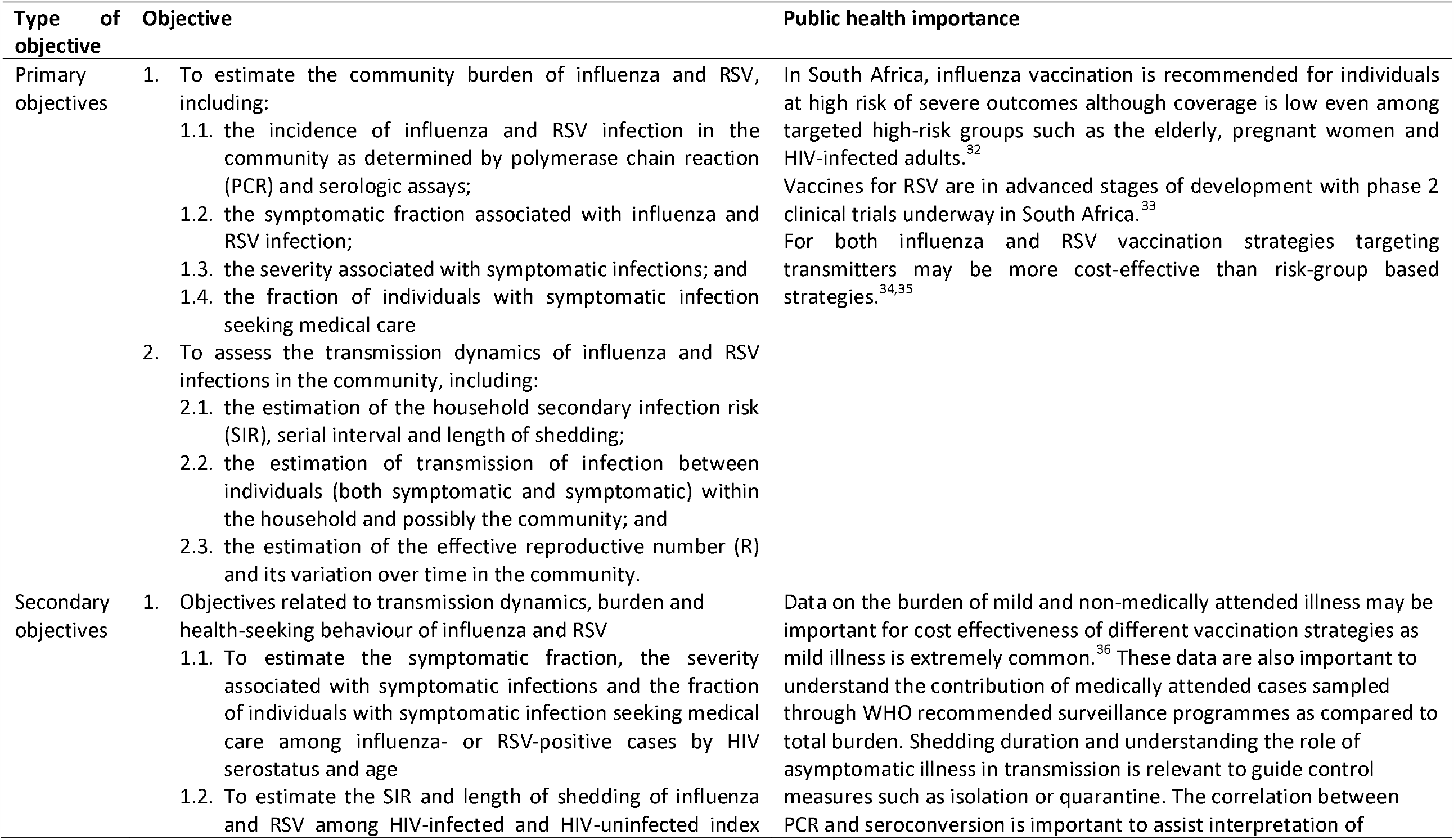
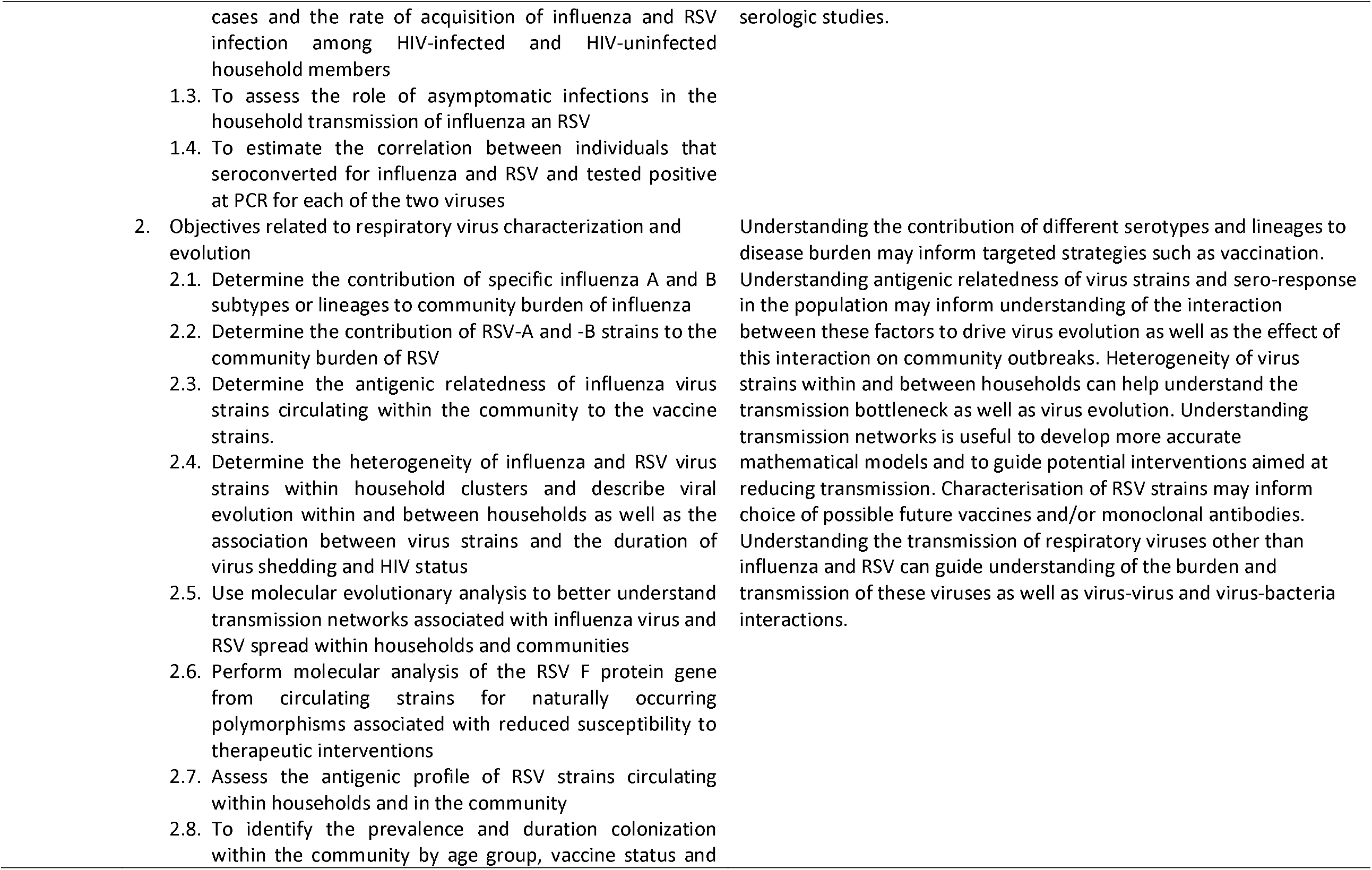
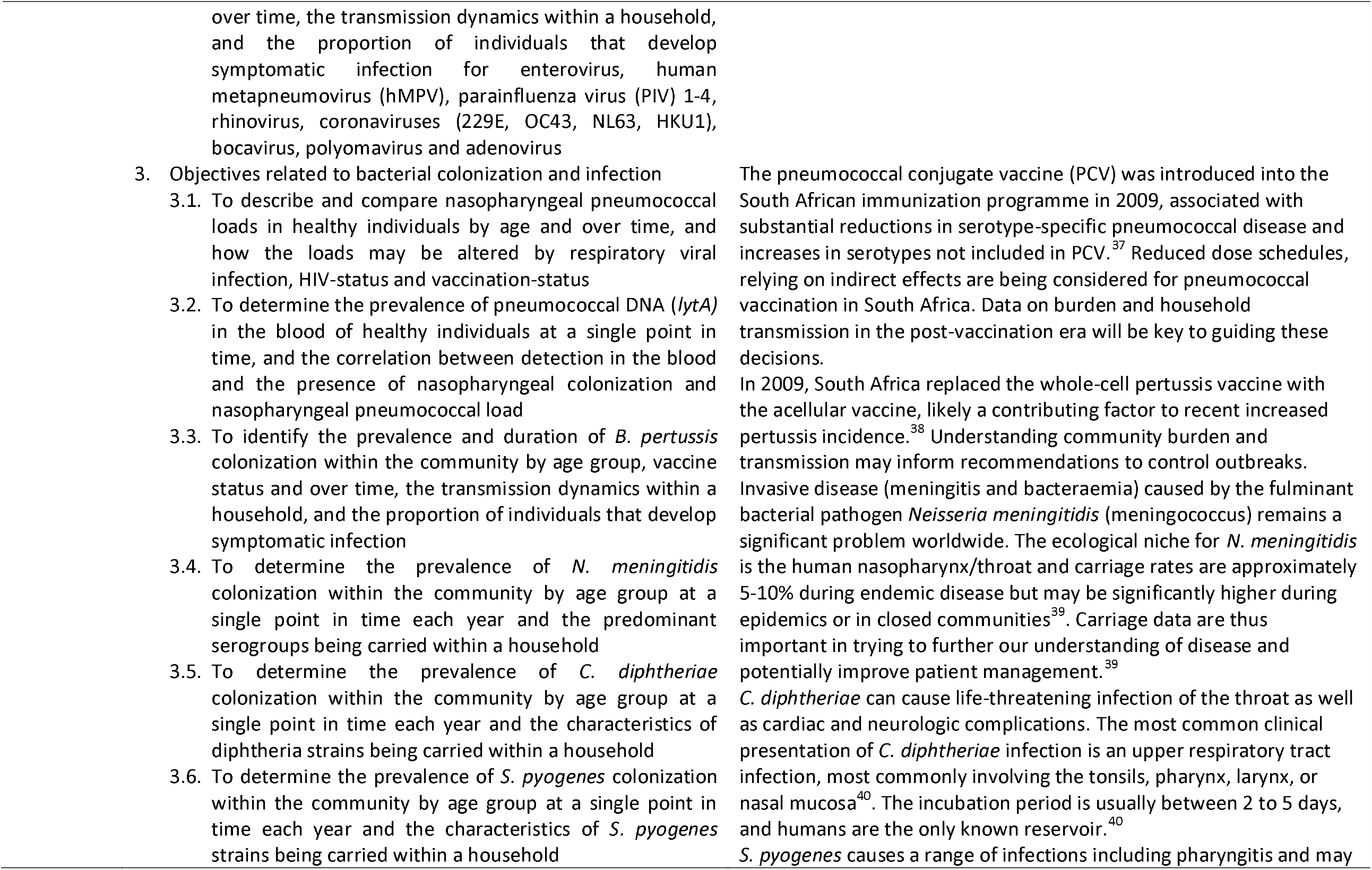
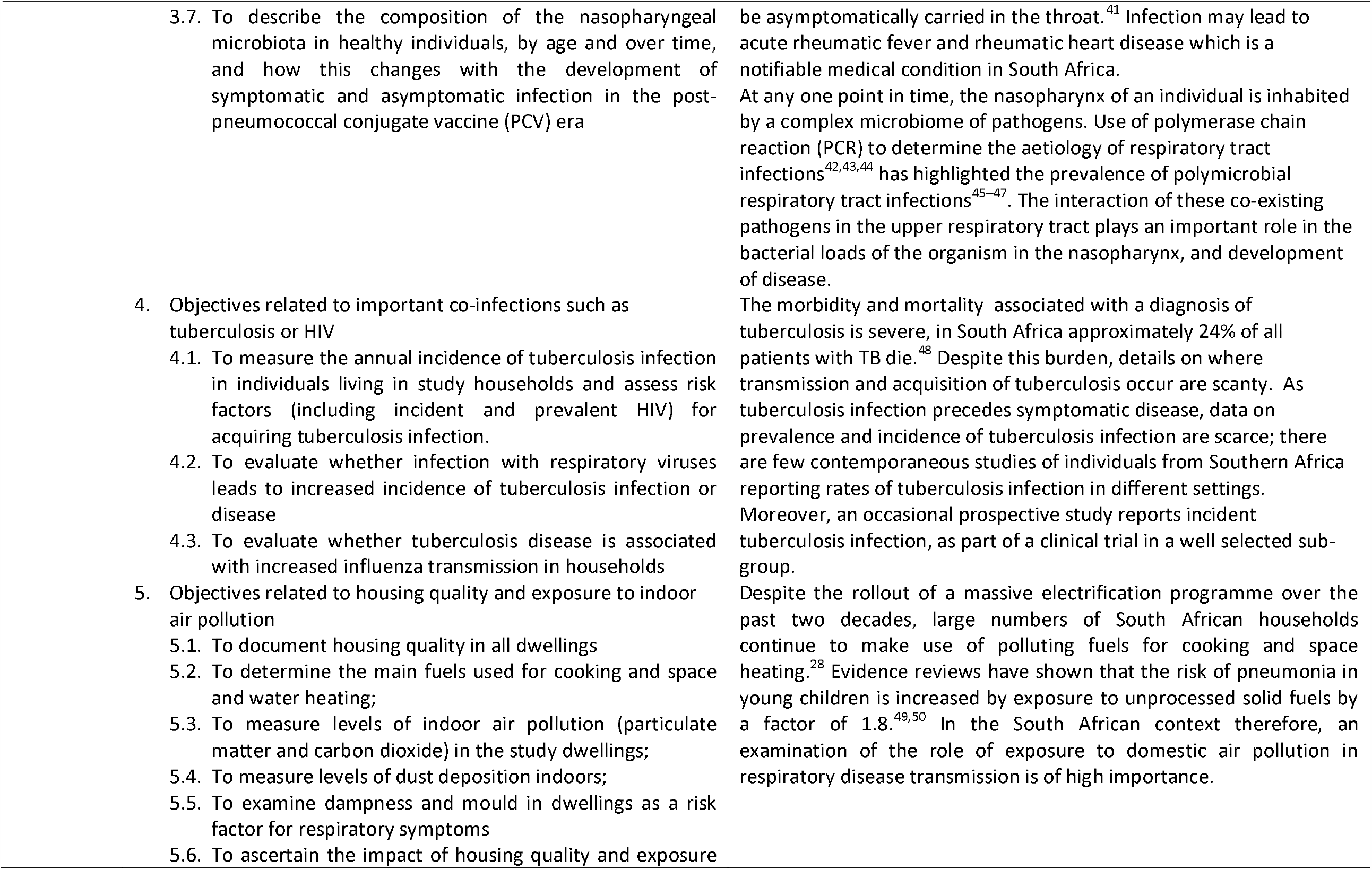
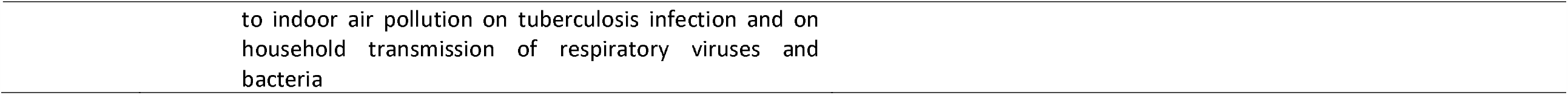
PHIRST study primary and secondary objectives and public health importance of questions

**Supplementary table 2:**
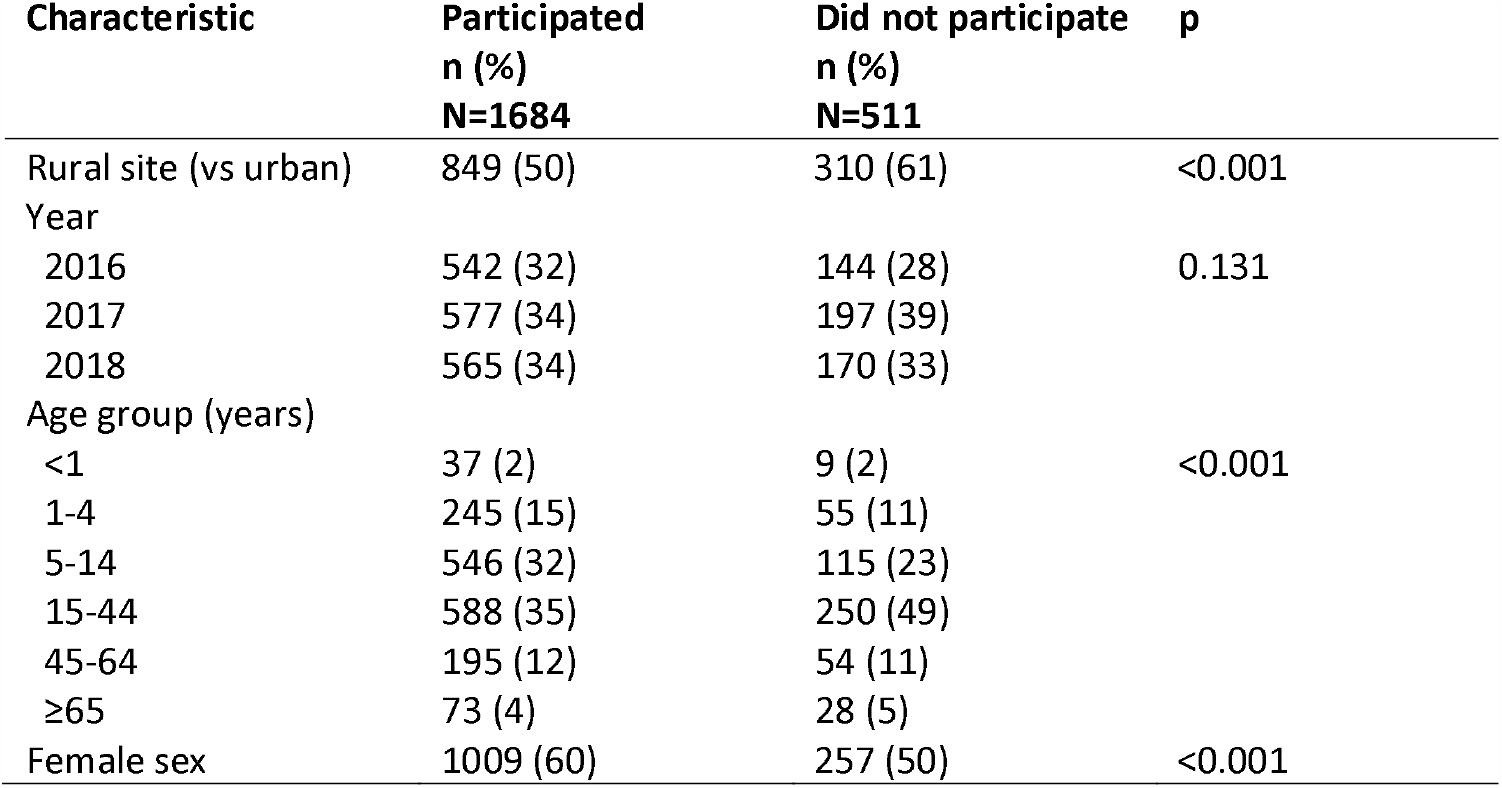
Comparison of characteristics of individuals who participated in the study compared to those who did not participate within included households, a rural and an urban site, South Africa, 2016-2018

**Supplementary table 3:**
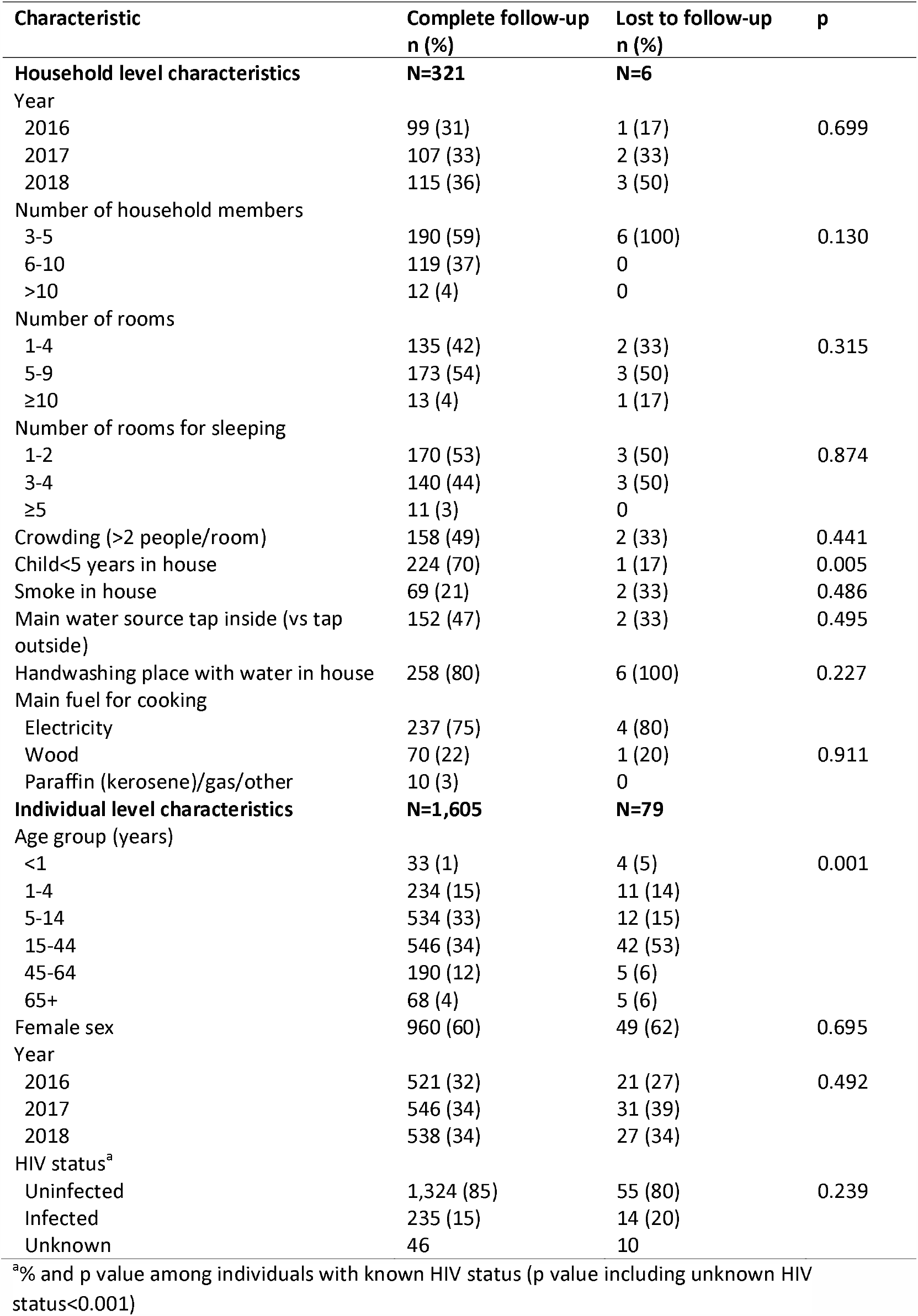
Comparison of individual and household-level characteristics of individuals lost to follow-up for twice-weekly visits to those with complete follow-up, a rural and an urban site, South Africa, 2016-2018

| Characteristic | Complete follow-up<br>n (%) | Lost to follow-up<br>n (%) | p |
| --- | --- | --- | --- |
| **Household level characteristics** | **N=321** | **N=6** |  |
| Year |  |  |  |
| 2016 | 99 (31) | 1 (17) | 0.699 |
| 2017 | 107 (33) | 2 (33) |  |
| 2018 | 115 (36) | 3 (50) |  |
| Number of household members |  |  |  |
| 3-5 | 190 (59) | 6 (100) | 0.130 |
| 6-10 | 119 (37) | 0 |  |
| >10 | 12 (4) | 0 |  |
| Number of rooms |  |  |  |
| 1-4 | 135 (42) | 2 (33) | 0.315 |
| 5-9 | 173 (54) | 3 (50) |  |
| ≥10 | 13 (4) | 1 (17) |  |
| Number of rooms for sleeping |  |  |  |
| 1-2 | 170 (53) | 3 (50) | 0.874 |
| 3-4 | 140 (44) | 3 (50) |  |
| ≥5 | 11 (3) | 0 |  |
| Crowding (>2 people/room) | 158 (49) | 2 (33) | 0.441 |
| Child<5 years in house | 224 (70) | 1 (17) | 0.005 |
| Smoke in house | 69 (21) | 2 (33) | 0.486 |
| Main water source tap inside (vs tap outside) | 152 (47) | 2 (33) | 0.495 |
| Handwashing place with water in house | 258 (80) | 6 (100) | 0.227 |
| Main fuel for cooking |  |  |  |
| Electricity | 237 (75) | 4 (80) | 0.911 |
| Wood | 70 (22) | 1 (20) |  |
| Paraffin (kerosene)/gas/other | 10 (3) | 0 |  |
| **Individual level characteristics** | **N=1,605** | **N=79** |  |
| Age group (years) |  |  |  |
| <1 | 33 (1) | 4 (5) | 0.001 |
| 1-4 | 234 (15) | 11 (14) |  |
| 5-14 | 534 (33) | 12 (15) |  |
| 15-44 | 546 (34) | 42 (53) |  |
| 45-64 | 190 (12) | 5 (6) |  |
| 65+ | 68 (4) | 5 (6) |  |
| Female sex | 960 (60) | 49 (62) | 0.695 |
| Year |  |  |  |
| 2016 | 521 (32) | 21 (27) | 0.492 |
| 2017 | 546 (34) | 31 (39) |  |
| 2018 | 538 (34) | 27 (34) |  |
| HIV status<sup>a</sup> |  |  |  |
| Uninfected | 1,324 (85) | 55 (80) | 0.239 |
| Infected | 235 (15) | 14 (20) |  |
| Unknown | 46 | 10 |  |

<sup>a</sup>% and p value among individuals with known HIV status (p value including unknown HIV status<0.001)

**Supplementary Figure 1:**
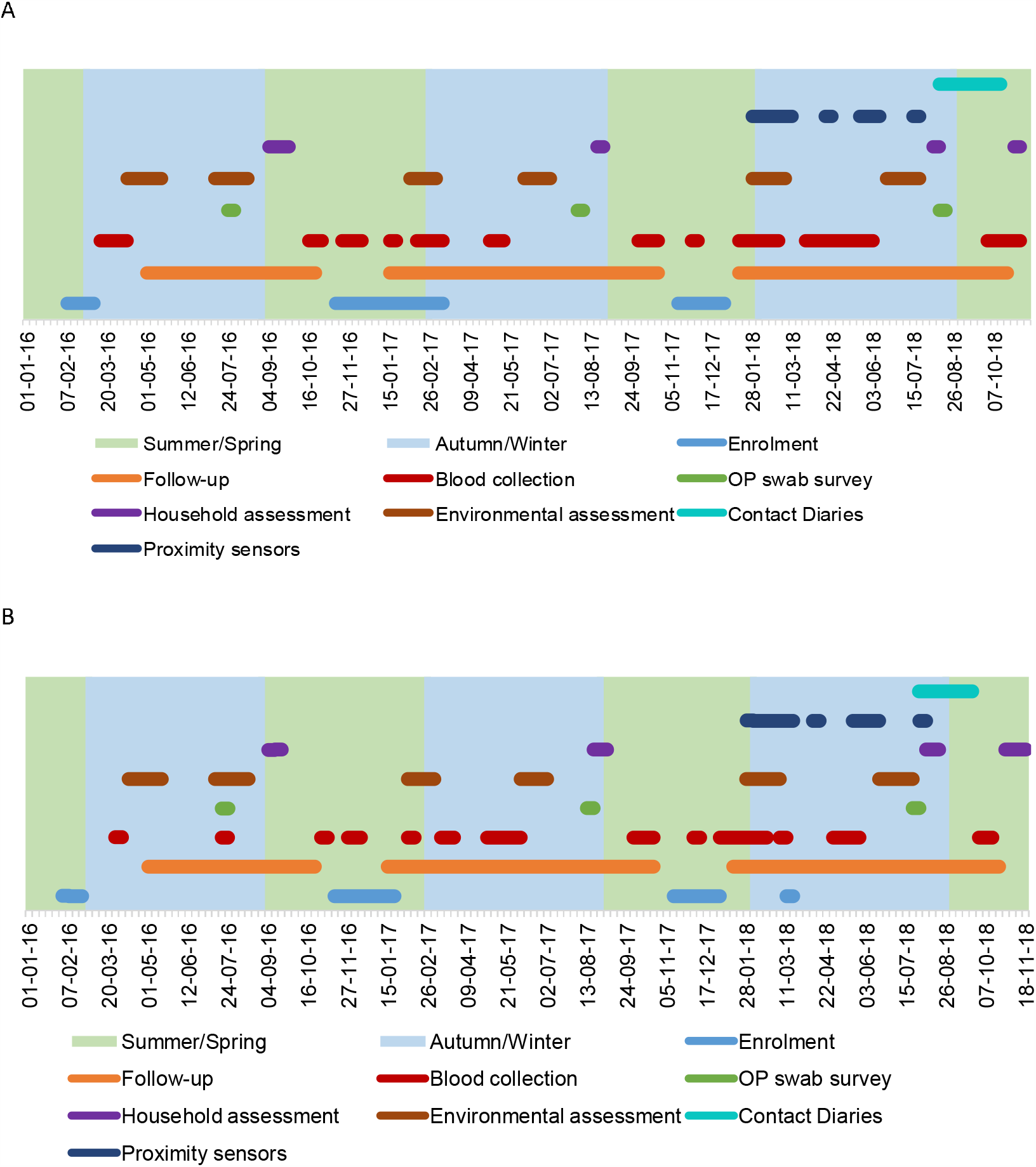
Timing of study procedures in relation to enrolment, follow up and season, a rural and an urban site, South Africa, 2016-2018 a)rural site b) urban site

